# Mortality after surgery with SARS-CoV-2 infection in England: A population-wide epidemiological study

**DOI:** 10.1101/2021.02.17.21251928

**Authors:** T. E. F. Abbott, A. J. Fowler, T. D. Dobbs, J. Gibson, T. Shahid, P. Dias, A. Akbari, I. S. Whitaker, R. M. Pearse

## Abstract

**Objectives:** To confirm the incidence of perioperative SARS-CoV-2 infection and associated mortality after surgery.

**Design and setting:** Analysis of routine electronic health record data from National Health Service (NHS) hospitals in England.

**Methods:** We extracted data from Hospital Episode Statistics in England describing adult patients undergoing surgery between 1^st^ January 2020 and 31^st^ October 2020. The exposure was SARS-CoV-2 infection defined by ICD-10 codes. The primary outcome measure was 90-day in-hospital mortality. Data were analysed using multivariable logistic regression adjusted for age, sex, Charlson co-morbidity index, index of multiple deprivation, presence of cancer, surgical procedure type and admission acuity. Results are presented as n (%) and odds ratios (OR) with 95% confidence intervals.

**Results:** We identified 1,972,153 patients undergoing surgery of whom 11,940 (0.6%) had SARS-CoV-2. In total, 19,100 (1.0%) patients died in hospital. SARS-CoV-2 infection was associated with a much greater risk of death (SARS-CoV-2: 2,618/11,940 [21.9%] vs No SARS-CoV-2: 16,482/1,960,213 [0.8%]; OR: 5.8 [5.5 – 6.1]; p<0.001). Amongst patients undergoing elective surgery 1,030/1,374,985 (0.1%) had SARS-CoV-2 of whom 83/1,030 (8.1%) died, compared with 1,092/1,373,955 (0.1%) patients without SARS-CoV-2 (OR: 29.0 [22.5 −37.3]; p<0.001). Amongst patients undergoing emergency surgery 9,742/437,891 (2.2%) patients had SARS-CoV-2, of whom 2,466/9,742 (25.3%) died compared with 14,817/428,149 (3.5%) patients without SARS-CoV-2 (OR: 5.7 [5.4 – 6.0]; p<0.001).

**Conclusions:** The low incidence of SARS-CoV-2 infection in NHS surgical pathways suggests current infection prevention and control policies are highly effective. However, the high mortality amongst patients with SARS-CoV-2 suggests these precautions cannot be safely relaxed.

**Summary boxes:** *What is already known on this topic:*

- High mortality rates have been reported amongst surgical patients who develop COVID-19 but we don’t know how this compares to the concurrent surgical population unaffected by COVID-19.
- Strict infection prevention and control procedures have substantially reduced the capacity of surgical treatment pathways in many hospitals.
- The very large backlog in delayed and cancelled surgical procedures is a growing public health concern.

*What this study adds:*

- Fewer than 1 in 100 surgical patients are affected by COVID-19 in the English National Health Service.
- Elective surgical patients who do develop COVID-19 are 30 times more likely to die while in hospital.
- Infection prevention and control procedures in NHS surgical pathways are highly effective but cannot be safely relaxed.

## Introduction

Surgery is an essential treatment modality with more than 330 million surgical procedures performed worldwide every year.^1-3^ However, the COVID-19 pandemic has led to substantial reductions in the volume of surgery performed.^4,5^ Recent estimates suggest that half of the 4.5 million expected surgical procedures in the English National Health Service (NHS) were cancelled or postponed in 2020.^4^ This is partly driven by reallocation of resources to care for patients with COVID-19, but also by strict infection prevention and control procedures implemented to prevent patients becoming infected with SARS-CoV-2 whilst in hospital.^6^ Patient self-isolation before surgery, pre-operative testing for SARS-CoV-2, creation of separate ‘green zones’ in hospitals to isolate non-infected patients, and personal protective equipment procedures all contribute to reductions in the efficiency of surgical care pathways, and substantial reductions in the volume of patients treated.^7^

Early reports have suggested that patients with SARS-CoV-2 infection who undergo surgery are at much greater risk of postoperative pulmonary complications and death.^8,9^ The COVIDSurg collaborative undertook a large international study of outcomes for surgical patients infected with SARS-CoV-2 and reported mortality rates as high as 24% at the peak of the pandemic.^8^ However, more recent data suggest the mortality risk for surgical patients with SARS-CoV-2 may be lower than originally thought,^9^ especially for minor surgery in younger patients.^10^ It remains unclear how SARS-CoV-2 infection affects outcomes after surgery, and whether high mortality rates reported in some studies relate to a change in surgical casemix during the pandemic, or to SARS-CoV-2 infection itself. At present there are no large studies describing surgical outcomes for contemporaneous patient groups with and without SARS-CoV-2 infection. Given the extensive disruption to surgical services and the likely excess mortality due to untreated cancer and other surgical diseases, the NHS is under significant pressure to relax infection prevention and control procedures to increase the volume of patients who can undergo surgery.

We used routine NHS electronic health record data to report the rate of SARS-CoV-2 infection amongst surgical patients during the pandemic, and the associated mortality and hospital stay.

## Methods

### Study design

Population-wide epidemiological study using routinely collected electronic health record data.

### Setting

NHS hospitals in England.

### Participants

All patients aged 18 years or older who underwent a surgical procedure between 1^st^ January and 31^st^ October 2020.

### Data source

We used pseudonymised record level Hospital Episode Statistics (HES) to identify eligible patients. This data source provides detailed data describing every episode of hospital care in England. Surgical procedures were identified using Office for Population Censuses Surveys version 4 (OPCS4) codes.^2,4^ Individuals entered the cohort on the date of their first operative procedure and were followed until the point of hospital discharge. Hospital discharge date was determined based on the discharge from continuous in-patient spells, which were constructed by mapping continuous in-patient episodes.^11^ For patients remaining in hospital, their discharge rate was right censored to the 31^st^ October 2020. The analysis was approved by the Health Research Authority (20/HRA/3121) and the NHS Digital Independent Group Advising on the Release of Data (DARS-NIC-375669-J7M7F).

### Exposure

The exposure of interest was a diagnosis of SARS-CoV-2 viral infection defined using the following ICD-10 codes: U07.1 (Virus identified), U07.2 (Virus not identified) or B97.2 (Other coronavirus as the cause of diseases classified elsewhere). Patients with SARS-CoV-2 were categorised as symptomatic if they had ICD-10 codes for concomitant respiratory illness or OPCS-4 codes indicating respiratory support (Supplementary table 1). Patients with SARS-CoV-2 were categorised as not symptomatic if there were no associated ICD-10 codes for respiratory illness or OPCS-4 codes for respiratory support. We divided the exposure into three time points: Preoperative if SARS-CoV-2 codes were first recorded in an episode that finished with the 30 days prior to the index operation; Perioperative if a code was first recorded in the same episode as the index operation; and Postoperative if a code was first recorded in a subsequent healthcare episode prior to discharge within 30 days after the index operation.

### Outcomes

The primary outcome was in-hospital death, censored at 90 days for patients remaining in hospital beyond this point. The secondary outcome was length of hospital stay, calculated as the number of days between initial operation date and discharge date of their continuous in-patient spell, censored at 90 days.

### Derivation of variables

Age was defined as that recorded on the start date of the hospital episode including the first operative procedure. Charlson co-morbidity index was derived according to the Royal College of Surgeons mapping which includes a one-year look back file to capture prior admissions.^12^ Classification of the type of surgical procedure was based on the first operative code.^4^ Where multiple operative codes were associated with surgery on a single day, the highest ranked code was considered the principal surgical procedure. Socioeconomic deprivation was defined according to lower super-output areas using the Index of Multiple Deprivation (IMD).^13^

### Statistical methods

The incidence of SARS-CoV-2 infection among patients undergoing surgery was calculated by dividing the number of patients with an ICD-10 code indicating SARS-CoV-2 infection by the total number of patients in the cohort. Age-adjusted incidence was calculated using deciles of age.^14^ To test for associations between SARS-CoV-2 infection and in-hospital mortality we used multivariable logistic regression analysis including the following covariates: age, sex, Charlson co-morbidity index, IMD, presence of cancer, surgical procedure type and admission acuity.^15-18^ Results are presented as n (%), mean (SD), median (IQR) or as odds ratios (OR) with 95% confidence intervals and p-values. The threshold for statistical significance was p<0.05. We undertook two pre-specified sub-group analyses. First, we assessed the risk of mortality among patients with SARS-CoV-2 infection stratified by urgency of surgery (elective surgery or emergency surgery). Second, we compared the risk of mortality among patients with SARS-CoV-2 infection between patients with and without respiratory symptoms. Analyses were conducted using Python (version 3.7.5) and graphs made in R (V4.0.2, R-project, Vienna).

### Sensitivity analysis using laboratory data from NHS Wales

To check the completeness of ICD-10 coding for a positive SARS-CoV-2 diagnosis, we compared reported cases of SARS-CoV-2 infection using ICD-10 codes to the integrated central laboratory records system describing all SARS-CoV-2 test results in Wales, which includes results of tests conducted for Welsh residents from both NHS and private laboratories. This analysis used pseudonymised record level Patient Episode Database in Wales (PEDW) to identify eligible patients. The project was approved by the Secure Anonymised Information Linkage Databank (SAIL) independent Information Governance Review Panel (Project number: 0911).^19^

### Sample size calculation

This was a population-wide cohort study including all patients that underwent surgery in England during the study period. A sample size of 2 million patients, with an allocation ratio of 0.01, and an alpha of 0.05 would give a 100% power to detect a 10% difference in the relative risk of mortality among patients with and without SARS-CoV-2 infection.

## Results

### Participants

There were 3,813,678 surgical procedures among 2,159,952 patients in England between 1^st^ January and 31^st^ October 2020. After predefined exclusions, 1,972,153 patients undergoing were included in the primary analysis (Figure 1). Demographic data are presented in Table 1. Data are presented according to surgical specialty in Table 2.

**Table 1.** Patient characteristics. Data are presented as mean (SD), median (IQR) and n(%)Numbers are presented as n (%) unless otherwise stated. *Numbers suppressed to maintain patient anonymity.

|  | All patients | SARS-CoV-2 status |  |  |  |  |
| --- | --- | --- | --- | --- | --- | --- |
|  |  | None | Preoperative | Perioperative | Postoperative | Any time |
| N | 1972153 | 1960213 (99.4%) | 3289 (0.2%) | 7514 (0.4%) | 1137 (0.1%) | 11940 (0.6%) |
| **Age** |  |  |  |  |  |  |
| Mean (SD) | 58 (20) | 58 (20) | 63 (17) | 64 (20) | 74 (15.7) | 64 (19) |
| Median (IQR) | 60 (40 to 74) | 60 (40 to 74) | 63 (52 to 76) | 66 (51 to 80) | 78 (67 to 86) | 67 (52 to 80) |
| **Sex** |  |  |  |  |  |  |
| Male | 883520 (44.8%) | 877002 (99.3%) | 1981 (0.2%) | 3941 (0.4%) | 596 (0.1%) | 6518 (0.7%) |
| Female | 1088536 (55.2%) | 1083114 (99.5%) | 1308 (0.1%) | 3573 (0.3%) | 541 (0%) | 5422 (0.5%) |
| **Admission category** |  |  |  |  |  |  |
| Elective | 1374985 (69.7%) | 1373955 (99.9%) | 360 (0%) | 612 (0%) | 60 (0%) | 1030 (0.1%) |
| Emergency | 437891 (22.2%) | 428149 (97.8%) | 2563 (0.6%) | 6135 (1.4%) | 1044 (0.2%) | 9742 (2.2%) |
| Other | 16086 (0.8%) | 15604 (97%) | 258 (1.6%) | 192 (1.2%) | 35 (0.2%) | 482 (3%) |
| Maternity | 143191 (7.3%) | 142505 (99.5%) | 108 (0.1%) | 575 (0.4%) | *(*) | 686 (0.5%) |
| Day-case | 973687(49.4%) | 973477 (100%) | 150 (0%) | 60 (0%) | *(*) | 210 (0%) |
| **Charlson comorbidity index** |  |  |  |  |  |  |
| Score > 0 | 922669 (99%) | 2644 (0.3%) | 5387 (0.6%) | 980 (0.1%) | 9011 (1%) | 922669 (99%) |
| **Index of multiple deprivation** |  |  |  |  |  |  |
| 1 – most deprived | 390173 (19.8%) | 387000 (99.2%) | 898 (0.2%) | 2004 (0.5%) | 271 (0.1%) | 3173 (0.8%) |
| 2 | 390220 (19.8%) | 387637 (99.3%) | 742 (0.2%) | 1630 (0.4%) | 211 (0.1%) | 2583 (0.7%) |
| 3 | 390136 (19.8%) | 387903 (99.4%) | 618 (0.2%) | 1388 (0.4%) | 227 (0.1%) | 2233 (0.6%) |
| 4 | 390170 (19.8%) | 388231 (99.5%) | 521 (0.1%) | 1198 (0.3%) | 220 (0.1%) | 1939 (0.5%) |
| 5 – least deprived | 390138 (19.8%) | 388304 (99.5%) | 468 (0.1%) | 1171 (0.3%) | 195 (0%) | 1834 (0.5%) |

**Table 2.**
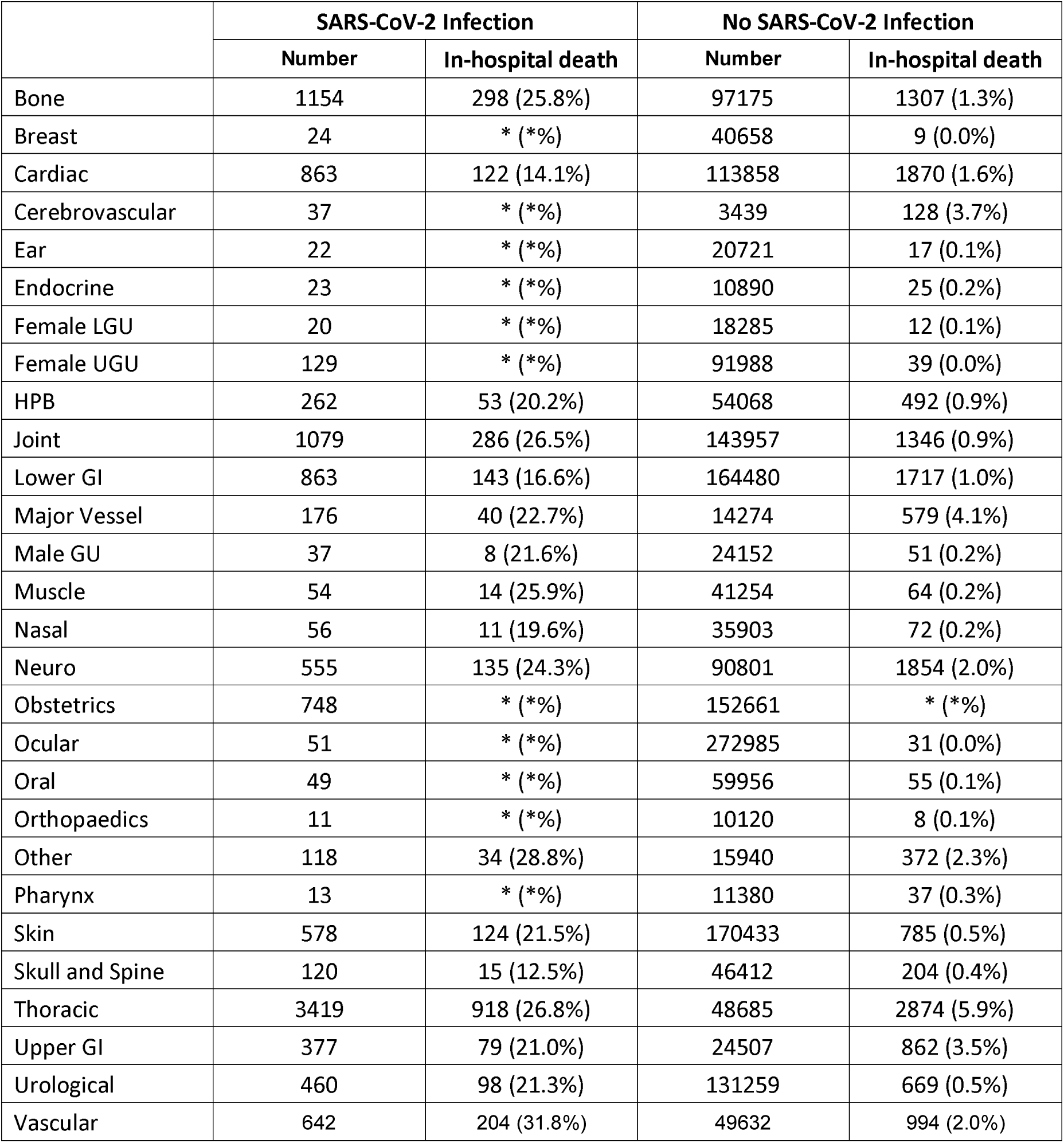
Risk of in-hospital mortality stratified by surgical specialty. Data presented as n (%); *Numbers suppressed to maintain patient anonymity. Risk of in-hospital mortality stratified by surgical specialty. Numbers are presented as n. *numbers suppressed. Organ donation removed given low numbers. LGU; Lower genitourinary, UGU; Upper genitourinary, HPB; hepatopancreatobiliary, GI; gastrointestinal.

|  | SARS-CoV-2 Infection |  | No SARS-CoV-2 Infection |  |
| --- | --- | --- | --- | --- |
|  | Number | In-hospital death | Number | In-hospital death |
| Bone | 1154 | 298 (25.8%) | 97175 | 1307 (1.3%) |
| Breast | 24 | * (%) | 40658 | 9 (0.0%) |
| Cardiac | 863 | 122 (14.1%) | 113858 | 1870 (1.6%) |
| Cerebrovascular | 37 | * (%) | 3439 | 128 (3.7%) |
| Ear | 22 | * (%) | 20721 | 17 (0.1%) |
| Endocrine | 23 | * (%) | 10890 | 25 (0.2%) |
| Female LGU | 20 | * (%) | 18285 | 12 (0.1%) |
| Female UGU | 129 | * (%) | 91988 | 39 (0.0%) |
| HPB | 262 | 53 (20.2%) | 54068 | 492 (0.9%) |
| Joint | 1079 | 286 (26.5%) | 143957 | 1346 (0.9%) |
| Lower GI | 863 | 143 (16.6%) | 164480 | 1717 (1.0%) |
| Major Vessel | 176 | 40 (22.7%) | 14274 | 579 (4.1%) |
| Male GU | 37 | 8 (21.6%) | 24152 | 51 (0.2%) |
| Muscle | 54 | 14 (25.9%) | 41254 | 64 (0.2%) |
| Nasal | 56 | 11 (19.6%) | 35903 | 72 (0.2%) |
| Neuro | 555 | 135 (24.3%) | 90801 | 1854 (2.0%) |
| Obstetrics | 748 | * (%) | 152661 | * (%) |
| Ocular | 51 | * (%) | 272985 | 31 (0.0%) |
| Oral | 49 | * (%) | 59956 | 55 (0.1%) |
| Orthopaedics | 11 | * (%) | 10120 | 8 (0.1%) |
| Other | 118 | 34 (28.8%) | 15940 | 372 (2.3%) |
| Pharynx | 13 | * (%) | 11380 | 37 (0.3%) |
| Skin | 578 | 124 (21.5%) | 170433 | 785 (0.5%) |
| Skull and Spine | 120 | 15 (12.5%) | 46412 | 204 (0.4%) |
| Thoracic | 3419 | 918 (26.8%) | 48685 | 2874 (5.9%) |
| Upper GI | 377 | 79 (21.0%) | 24507 | 862 (3.5%) |
| Urological | 460 | 98 (21.3%) | 131259 | 669 (0.5%) |
| Vascular | 642 | 204 (31.8%) | 49632 | 994 (2.0%) |

**Figure 1.**
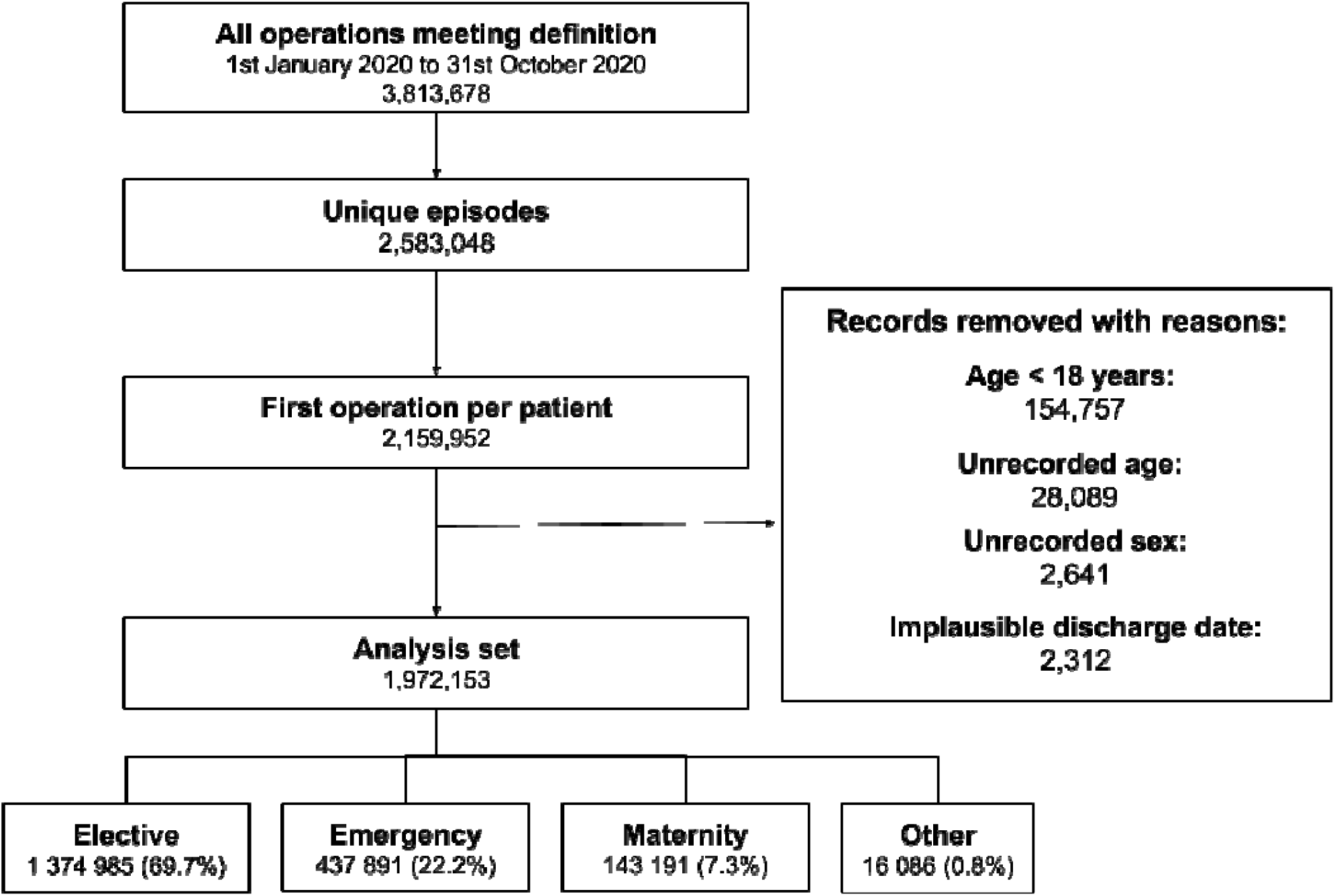
Inclusion of patients in the analysis.

### Incidence of SARS-CoV-2 among surgical patients

Among 1,972,153 adult patients who underwent surgery, 11,940 (0.6%) had SARS-CoV-2 (Table 1 and Figure 2). 8,151/12,482 (65.3%) surgical patients with SARS-CoV-2 infection had respiratory symptoms. The age-adjusted incidence of SARS-CoV-2 infection among surgical patients was 527 infections per 100,000 head of population (496-559).

**Figure 2.**
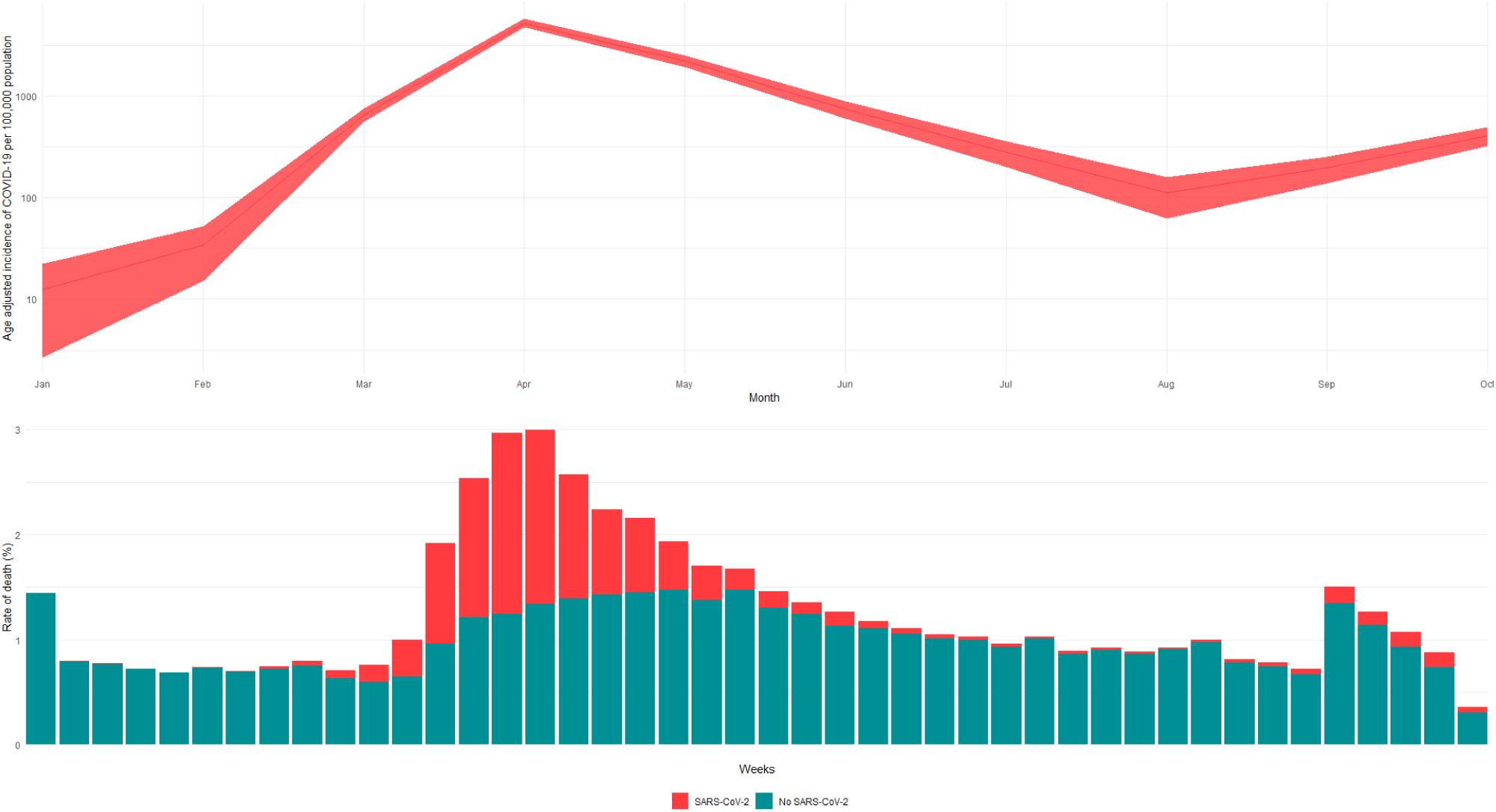
**Top panel: Age-adjusted incidence of SARS-CoV-2 infection amongst surgical patients, reported as the number of cases per 100,000 population (y axis on log scale). Bottom panel: Weekly rate of death, stratified by SARS-COV-2 infection rate.**

**Figure 3.**
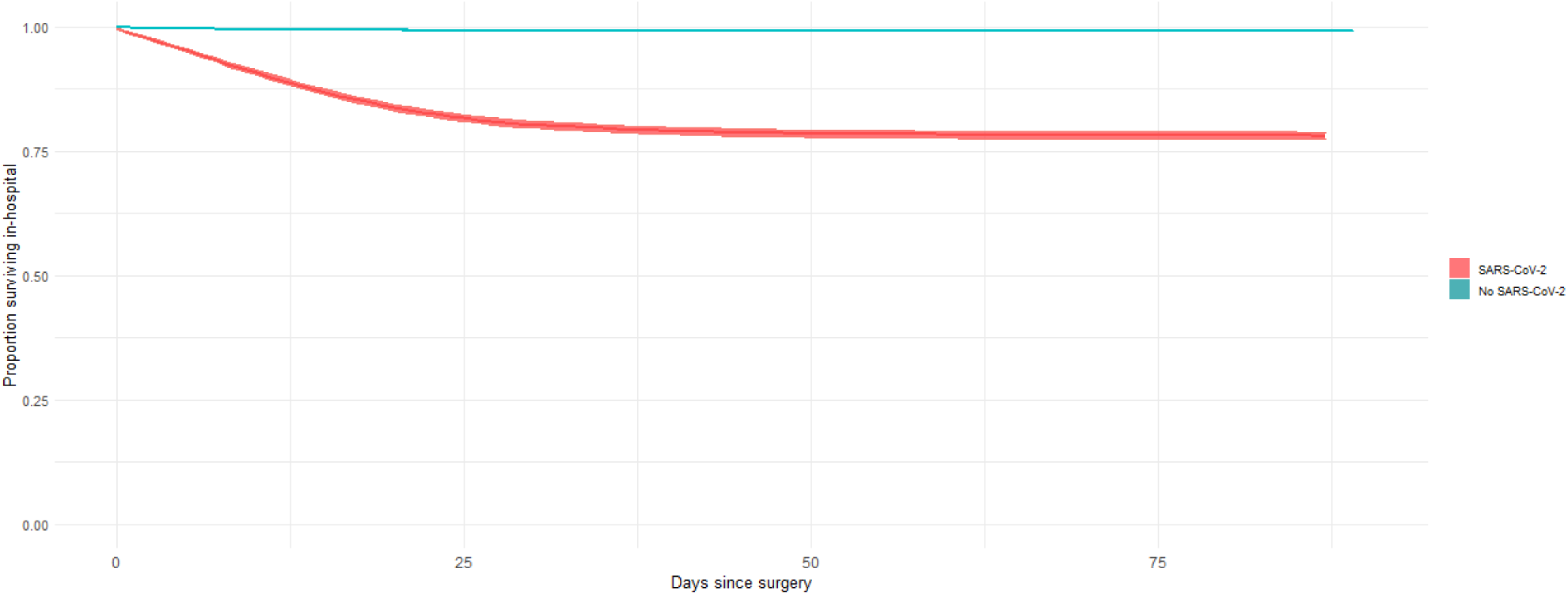
Kaplan Meier survival curves stratified by SARS-CoV-2 status.

### Patient outcomes

19,100/1,972,153 (1.0%) patients who underwent a surgical procedure died. 2,618/11,940 (21.9%) surgical patients with SARS-CoV-2 infection died compared to 16,482/1,960,213 (0.8%) patients without SARS-CoV-2 infection (Table 3). The adjusted odds of death among surgical patients with SARS-CoV-2 infection compared to surgical patients without infection is 5.8 (5.5-6.1; p<0.001) (Table 4). Length of hospital stay among patients with and without SARS-CoV-2 infection is presented in Table 3.

**Table 3.** Mortality and hospital length of stay, stratified by SARS-CoV-2 infection status and acuity of surgery. Data are presented as median (IQR) and n (%).

|  | No SARS-CoV-2 | SARS-CoV-2 Infection |  |  |  |
| --- | --- | --- | --- | --- | --- |
|  |  | Any | Preoperative | Perioperative | Postoperative |
| All patients |  |  |  |  |  |
| N | 1960213 | 11940 | 3289 | 7514 | 1137 |
| In-hospital death | 16482 (0.8%) | 2618 (21.9%) | 616 (18.7%) | 1607 (21.4%) | 395 (34.7%) |
| Length of hospital stay (days) | 0 (0 to 2) | 13 (5 to 27) | 11 (3 to 25) | 13 (4 to 28) | 22.0 (14 to 30) |
| Elective Surgery |  |  |  |  |  |
| N | 1373955 | 1030 | 360 | 612 | 58 |
| In-hospital death | 1092 (0.1%) | 83 (8.1%) | 11 (3.1%) | 59 (9.6%) | 13 (22.4%) |
| Length of hospital stay (days) | 0 (0 to 1) | 5 (1 to 15) | 1.0 (0 to 4) | 7 (2 to 18.2) | 23.5 (14 to 30) |
| Emergency Surgery |  |  |  |  |  |
| N | 428149 | 9742 | 2563 | 6135 | 1044 |
| In-hospital death | 14817 (3.5%) | 2466 (25.3%) | 575 (22.4%) | 1518 (24.7%) | 373 (35.7%) |
| Length of hospital stay (days) | 2 (1 to 8) | 16 (7 to 29) | 13 (5 to 27) | 15 (6 to 30) | 21 (14 to 29) |

**Table 4.** Multivariable logistic regression analysis for in-hospital mortality within 90 days. Patients with unspecified sex removed prior to analysis.LGU; Lower genitourinary, UGU; Upper genitourinary, HPB; hepatopancreatobiliary, GI; gastrointestinal.

|  | Odds ratio | Lower 95%CI | Upper 95%CI | p |
| --- | --- | --- | --- | --- |
| SARS-CoV-2 code present | 5.81 | 5.52 | 6.12 | <0.001 |
| Female compared to Male | 0.85 | 0.83 | 0.88 | <0.001 |
| Age in years | 1.03 | 1.03 | 1.03 | <0.001 |
| Cancer present | 1.02 | 0.99 | 1.06 | 0.22 |
| **Charlson co-morbidity score: reference = 0** |  |  |  |  |
| score 1 | 2.45 | 2.31 | 2.61 | <0.001 |
| score 2 | 3.93 | 3.7 | 4.19 | <0.001 |
| score 3 | 5.13 | 4.8 | 5.48 | <0.001 |
| score 4 | 6.44 | 5.98 | 6.93 | <0.001 |
| score 5 | 7.42 | 6.82 | 8.08 | <0.001 |
| score ≥ 6 | 8.19 | 7.43 | 9.03 | <0.001 |
| **Index of multiple deprivation: reference = quintile 5 (least deprived)** |  |  |  |  |
| quintile 1 (most deprived) | 0.96 | 0.92 | 1.01 | 0.08 |
| quintile 2 | 0.91 | 0.86 | 0.95 | <0.001 |
| quintile 3 | 0.89 | 0.85 | 0.93 | <0.001 |
| quintile 4 | 0.86 | 0.81 | 0.9 | <0.001 |
| **Admission category: reference = Elective** |  |  |  |  |
| Emergency | 24.07 | 22.62 | 25.61 | <0.001 |
| Maternity | 3.97 | 1.15 | 13.78 | 0.03 |
| Other | 17.82 | 16.05 | 19.79 | <0.001 |
| **Procedure grouping: reference = Bone** |  |  |  |  |
| Breast | 0.2 | 0.11 | 0.39 | <0.001 |
| Cardiac | 1.03 | 0.96 | 1.11 | 0.43 |
| Cerebrovascular | 2.62 | 2.17 | 3.17 | <0.001 |
| Ear | 0.39 | 0.26 | 0.6 | <0.001 |
| Endocrine | 1.52 | 1.03 | 2.26 | 0.04 |
| Female LGU | 0.47 | 0.28 | 0.8 | 0.01 |
| Female UGU | 0.4 | 0.3 | 0.55 | <0.001 |
| HPB | 1.59 | 1.43 | 1.76 | <0.001 |
| Joint | 1.13 | 1.05 | 1.22 | <0.001 |
| Lower GI | 1.8 | 1.68 | 1.93 | <0.001 |
| Major Vessel | 2.82 | 2.55 | 3.13 | <0.001 |
| Male GU | 0.76 | 0.58 | 1 | 0.05 |
| Muscle | 0.88 | 0.69 | 1.11 | 0.28 |
| Nasal | 0.45 | 0.36 | 0.56 | <0.001 |
| Neuro | 3.7 | 3.44 | 3.97 | <0.001 |
| Organ Donation | 22.61 | 6.97 | 73.33 | <0.001 |
| Obstetrics | 0.13 | 0.03 | 0.47 | <0.001 |
| Ocular | 0.09 | 0.07 | 0.13 | <0.001 |
| Oral | 0.63 | 0.48 | 0.82 | <0.001 |
| Orthopaedics | 0.4 | 0.2 | 0.82 | 0.01 |
| Other | 2.92 | 2.59 | 3.29 | <0.001 |
| Pharynx | 1.87 | 1.35 | 2.6 | <0.001 |
| Skin | 0.84 | 0.77 | 0.92 | <0.001 |
| Skull and Spine | 1.47 | 1.27 | 1.71 | <0.001 |
| Thoracic | 3.87 | 3.63 | 4.13 | <0.001 |
| Upper GI | 3.33 | 3.05 | 3.64 | <0.001 |
| Urological | 0.95 | 0.87 | 1.04 | 0.27 |
| Vascular | 1.78 | 1.64 | 1.93 | <0.001 |

### Urgency of surgery

Amongst patients undergoing elective surgery 1,030/1,373,995 (0.1%) had SARS-CoV-2 infection, of whom 83/1,030 (8.1%) died compared to 1,092/1,373,955 (0.1%) patients without SARS-CoV-2 infection (Table 3). Therefore, the incidence of death from SARS-CoV-2 infection among elective surgical patients was 83/1,373,995 (0.006% or 1 in 16556). The adjusted odds of death among patients undergoing elective surgery with SARS-CoV-2 around the time of surgery is 29.0 (22.5-37.3; p <0.001) (Supplementary table 2). Amongst patients undergoing emergency surgery 9,742/437,891 (2.2%) had SARS-CoV-2 infection, of whom 2,466/9,742 (25.3%) died compared with 14,817/428,149 (3.5%) patients without SARS-CoV-2 infection. The adjusted odds of death among patients undergoing emergency surgery with SARS-CoV-2 around the time of surgery is 5.7 (5.4-6.0; p<0.001) (Supplementary table 3).

### Symptomatic SARS-CoV-2 infection

Among patients with SARS-CoV-2 infection and respiratory symptoms 2,274/7,769 (29.3%) died compared to 344/4,171 (8.2%) patients with SARS-CoV-2 infection but no respiratory symptoms recorded (OR 3.7 [3.3–4.3]; p < 0.001) (Supplementary tables 4 and 5).

### Incidence of SARS-CoV-2 infection among surgical patients in Wales

We identified 86,866 patients undergoing their first surgical procedure in Wales between 1^st^ January and 31^st^ October 2020. Using ICD-10 codes, 730/86,866 (0.8%) surgical patients had SARS-CoV-2 infection. Adding positive SARS-CoV-2 polymerase chain reaction (PCR) test resulted in a marginally increased the incidence of 814/86,866 (0.9%). (Supplementary table 6).

## Discussion

The principal finding of this population-wide epidemiological study is that the national incidence of SARS-CoV-2 infection among NHS surgical patients in England is 0.6%. However, where perioperative SARS-CoV-2 infection does occur, it is associated with more than a five-fold increase in risk of in-hospital mortality. Meanwhile, the very low risk of in-hospital death (one in 1000) among patients undergoing elective surgery who have not been infected with SARS-CoV-2 suggests that infection prevention and control procedures to create COVID-19-free ‘green’ surgical pathways have been highly effective during the pandemic.^20,21^ Surgical patients with symptomatic SARS-CoV-2 infection were almost four times more likely to die compared to infected patients without respiratory symptoms.

The incidence of SARS-CoV-2 infection and subsequent deaths reported in this study represent the measured population incidences rather than an estimate from a population sample. Our findings are consistent with previous estimates of perioperative mortality associated with SARS-CoV-2 infection.^22-27^ The largest of these is the COVIDSurg international multicentre observational cohort study of 1,128 surgical patients with confirmed SARS-CoV-2 infection, which was conducted during the first quarter of 2020.^8^ It reported a 30-day mortality rate of 23.8%. However, the cohort consisted of only patients with confirmed SARS-CoV-2, so it was not possible to infer the relative risk of mortality associated with perioperative SARS-CoV-2 infection compared to a contemporaneous non-infected comparator. Our data contextualises these findings from early in the pandemic by reporting the population risk of mortality among surgical patients with SARS-CoV-2 in England using a cohort of patients with SARS-CoV-2 infection that is more than ten times larger. Since we included all patients undergoing surgery in England, we were able to compare the risk of mortality among patients with SARS-CoV-2 infection to patients without infection to provide an estimate of the relative risk of death associated with SARS-CoV-2 infection. This important finding, which until now was unknown, could inform shared decision making about surgical care of patients with SARS-CoV-2.

The volume of surgical activity during the pandemic was greatly reduced for three main reasons.^4-6,28,29^ First, the reduced capacity for elective surgery as a result of reallocation of staff and resources to the care of hospitalised patients with COVID-19 pneumonia.^30^ Second, the reorganisation of care pathways and the introduction of necessary, but onerous, infection control procedures have slowed down the delivery of care and patient throughput.^21,31^ Third, the reluctance of some clinicians to operate on patients at high-risk of complications should they suffer a nosocomial infection with SARS-CoV-2.^5,32^ This is coupled with a reluctance of some patients to undergo surgery due to fears of acquiring SARS-CoV-2 in hospital at the time of surgery. Our data confirm that the volume of surgical activity during 2020 is about 25% lower than expected compared to previous years.^1,2^ However, patients and clinicians should be reassured that the population incidence of SARS-CoV-2 among surgical patients is low and the overall risk of death from SARS-CoV-2 for a patient undergoing elective surgery in a ‘green’ pathway is less than one in 16500.^20^ The predicted delays to surgical treatments as a result of the pandemic are substantial, with an estimated five million cases outstanding by March 2021.^4^ Although the morbidity and mortality associated with cancelled or postponed surgery is unknown, it is clearly not desirable to have a large and rapidly expanding waiting list for treatments.^5,31^ Our data confirm that while it is possible to undertake surgery safely during the pandemic, the risk of mortality among surgical patients with SARS-CoV-2 is substantial and all efforts should be made to prevent nosocomial infection.

This study has several strengths. We included data from all patients undergoing surgery in England during the first wave of the COVID-19 pandemic in England, thus our results represent the true population incidence of SARS-CoV-2 infection and associated mortality. We were able to include data from almost two million surgical patients, which represents one of the largest observational cohorts of surgical patients during the COVID-19 pandemic. These data will be generalisable to other high-income countries. A major strength of our analysis is that we controlled for changing surgical casemix during the pandemic, as well as potential factors that could confound association between SARS-CoV-2 infection and mortality. We used the contemporaneous comparator of patients undergoing surgery during the pandemic who did not have SARS-CoV-2 infection. This allowed us to control for unexpected changes in surgical casemix associated with changes in population behaviour and healthcare delivery. We explored the potential for misclassification of SARS-CoV-2 status using a sample of patients from Wales with detailed SARS-CoV-2 testing data. The incidence of SARS-CoV-2 in this cohort was very similar to the overall incidence across the whole study, suggesting coding of SARS-CoV-2 using ICD-10 is accurate. Our analysis also has some limitations. It possible that our data underestimate the true incidence of SARS-CoV-2 infection among surgical patients, particularly amongst those undergoing day case procedures, which would lead to under-representation of asymptomatic patients who are less likely to die. However, we only included in-hospital deaths, and our findings will therefore underestimate the true mortality risk.

In conclusion, we have shown that the prevalence of SARS-CoV-2 infection among surgical patients is low and the risk of postoperative mortality among patients without SARS-CoV-2 infection is very small. However, where it occurs, SARS-CoV-2 infection among surgical patients is associated with a very high risk of death.

## Data Availability

The data used in this study are derived from two data sources. It is not possible to share the raw patient-level data provided by NHS Digital describing NHS patients in England. Regarding data from NHS patients in Wales, the data used are available in the SAIL Databank at Swansea University, Swansea, UK, but as restrictions apply they are not publicly available. All proposals to use SAIL data are subject to review by an independent Information Governance Review Panel (IGRP). Before any data can be accessed, approval must be given by the IGRP. The IGRP gives careful consideration to each project to ensure proper and appropriate use of SAIL data. When access has been granted, it is gained through a privacy protecting safe haven and remote access system referred to as the SAIL Gateway. SAIL has established an application process to be followed by anyone who would like to access data via SAIL at https://www.saildatabank.com/application-process.

## Contributors

TA, AF, TD, RP were responsible for study design. AF, GD, and AA were responsible for data collection. AF, TA and JG were responsible for data analysis. AF, TD, RP and TA were responsible for data interpretation. TA and RP wrote the first draft of the manuscript. All authors revised the manuscript for important intellectual content and approved the final version. AJF and TA had full access to the data and act as guarantors.

## Competing interest statement

All authors have completed the Unified Competing Interest form and declare: AJF holds a National Institute for Health Research Doctoral Research fellowship (DRF-2018-11-ST2-062). TDD reports funding from the Welsh Clinical Academic Training (WCAT) Fellowship. IW reports active grants from the American Association of Plastic Surgeons and the European Association of Plastic Surgeons; is an editor for Frontiers of Surgery, associate editor for the Annals of Plastic Surgery, editorial board of BMC Medicine and numerous other editorial board roles. RP has received honoraria and/or research grants from Edwards Lifesciences, Intersurgical and GlaxoSmithkline within the last five years and holds editorial roles with the British Journal of Anaesthesia, the British Journal of Surgery and BMJ Quality and Safety. TA is a member of the associate editorial board of the British Journal of Anaesthesia and has received consultancy fees from MSD unrelated to this work. All other authors report no financial relationships with any organisations that might have an interest in the submitted work in the previous three years, no other relationships or activities that could appear to have influenced the submitted work.

## Transparency declaration

TA affirms that the manuscript is an honest, accurate, and transparent account of the study being reported; that no important aspects of the study have been omitted; and that any discrepancies from the study as planned have been explained.

## Role of the funding source

This study was funded by a grant from Barts Charity. The Welsh data source was supported by Health Data Research UK, which receives its funding from HDR UK Ltd (NIWA1) funded by the UK Medical Research Council, Engineering and Physical Sciences Research Council, Economic and Social Research Council, Department of Health and Social Care (England), Chief Scientist Office of the Scottish Government Health and Social Care Directorates, Health and Social Care Research and Development Division (Welsh Government), Public Health Agency (Northern Ireland), British Heart Foundation (BHF) and the Wellcome Trust. The funding sources had no role in the study design, data collection, analysis, interpretation, or writing the report.

## Acknowledgement

This work uses data provided by patients and collected by the NHS as part of their care and support. We would like to acknowledge all data providers who make anonymised data available for research and the collaborative partnership that enabled acquisition and access to the de-identified data, which led to this output. The collaboration was led by the Swansea University Health Data Research UK team under the direction of the Welsh Government Technical Advisory Cell (TAC) and includes the following groups and organisations: the Secure Anonymised Information Linkage (SAIL) Databank, Administrative Data Research (ADR) Wales, NHS Wales Informatics Service (NWIS), Public Health Wales, NHS Shared Services and the Welsh Ambulance Service Trust (WAST). All research conducted has been completed under the permission and approval of the SAIL independent Information Governance Review Panel (IGRP) project number 0911.

## Patient and public involvement

A patient representative was consulted in the design of this study.

## Dissemination declaration

The results of this study have been shared with representatives from NHS England prior to publication, in order to inform patient care.

## Licence

The Corresponding Author has the right to grant on behalf of all authors and does grant on behalf of all authors, a worldwide licence (http://www.bmj.com/sites/default/files/BMJ%20Author%20Licence%20March%202013.do c) to the Publishers and its licensees in perpetuity, in all forms, formats and media (whether known now or created in the future), to i) publish, reproduce, distribute, display and store the Contribution, ii) translate the Contribution into other languages, create adaptations, reprints, include within collections and create summaries, extracts and/or, abstracts of the Contribution and convert or allow conversion into any format including without limitation audio, iii) create any other derivative work(s) based in whole or part on the on the Contribution, iv) to exploit all subsidiary rights to exploit all subsidiary rights that currently exist or as may exist in the future in the Contribution, v) the inclusion of electronic links from the Contribution to third party material where-ever it may be located; and, vi) licence any third party to do any or all of the above. All research articles will be made available on an open access basis (with authors being asked to pay an open access fee— see http://www.bmj.com/about-bmj/resources-authors/forms-policies-and-checklists/copyright-open-access-and-permission-reuse). The terms of such open access shall be governed by a Creative Commons licence—details as to which Creative Commons licence will apply to the research article are set out in our worldwide licence referred to above.

## Notes

### Author Declarations

The analysis was approved by the Health Research Authority (20/HRA/3121) and the NHS Digital Independent Group Advising on the Release of Data (DARS-NIC-375669-J7M7F).

