## Supplementary appendices for "Mortality after surgery with SARS-CoV-2 infection in England: A population-wide epidemiological study"

**T. E. F. Abbott and A. J. Fowler, T. D. Dobbs, J. Gibson, T. Shahid, P. Dias, A. Akbari, I. S. Whitaker, R. M. Pearse.**

* Joint first authors

**SUPPLEMENTARY FILE**

| **Page** | **Item** |
| --- | --- |
| 3 | Supplementary table 1:  **Codes defining SARS-CoV-2 status and symptomatic status** |
| 4 | Supplementary table 2:  **Multivariable logistic regression output: elective surgery** |
| 5 | Supplementary table 3:  **Multivariable logistic regression output: emergency surgery** |
| 6 | Supplementary table 4:  **Symptom status and outcomes among patients with SARS-CoV-2** |
| 7 | Supplementary table 5:  **Multivariable logistic regression output: symptom status** |
| 8 | Supplementary table 6:  **Incidence of SARS-CoV-2 measured by PCR or ICD-10 codes in Wales** |
| 9 | Supplementary table 7:  **OPCS codes excluded from the definition of surgery** |
| 10 | Statistical analysis plan. |

**Contents page**

| **Description** | **Code** | **SARS** | **SYMP** |
| --- | --- | --- | --- |
| SARS-CoV-2/COVID-19 (ICD10) | U071, U072 |  |  |
| Other coronavirus (ICD10) | B972 |  |  |
| Respiratory illness (ICD10) | J40, J208, J22, J988, J80, J128, J18, J16, J17 |  |  |
| Respiratory symptoms (ICD10) | R05, R06, R509, |  |  |
| Respiratory support (OPCS) | E851, E852, X581, X878, X879, X528, X529 |  |  |

**Supplementary table 1. Codes used for identification of relevant exposures.** SARS; record assigned SARS-CoV-2 status if any codes from green, or codes from both cells in orange present. SYMP; record assigned symptomatic if any of the codes from the green cells were present. OPCS; Office for Population Censuses Surveys version 4. ICD-10; International Statistical Classification of Diseases and Related Health Problems, 10^th^ Revision.

| **Variable** | **Odds ratio** | **Lower 95%CI** | **Upper 95%CI** | **p** |
| --- | --- | --- | --- | --- |
| SARS-CoV-2 code present | 28.97 | 22.49 | 37.34 | 0 |
| Female vs Male | 0.83 | 22.5 | 37.3 | 0 |
| Age | 1.03 | 1.03 | 1.04 | 0 |
| Cancer present | 1.32 | 1.14 | 1.53 | 0 |
| **Charlson co-morbidity score: vs.0** | | | | |
| Score 1 | 5.39 | 3.86 | 7.53 | 0.00 |
| Score 2 | 12.42 | 8.89 | 17.36 | 0.00 |
| Score 3 | 21.01 | 14.88 | 29.67 | 0.00 |
| Score 4 | 27.96 | 19.43 | 40.25 | 0.00 |
| Score 5 | 43.27 | 29.28 | 63.94 | 0.00 |
| Score >= 6 | 46.46 | 29.96 | 72.02 | 0.00 |
| **Index of multiple deprivation: vs quintile 5 (least deprived)** | | | | |
| Quintile 1 (most deprived) | 0.98 | 0.81 | 1.17 | 0.79 |
| Quintile 2 | 0.88 | 0.73 | 1.06 | 0.18 |
| Quintile 3 | 0.85 | 0.70 | 1.03 | 0.09 |
| Quintile 4 | 1.06 | 0.88 | 1.27 | 0.54 |
| **Procedure grouping: vs. Bone** | | | | |
| Breast | 0.06 | 0.01 | 0.29 | 0.00 |
| Cardiac | 2.33 | 1.26 | 4.31 | 0.01 |
| Ear | 0.08 | 0.01 | 0.58 | 0.01 |
| Endocrine | 0.41 | 0.09 | 1.84 | 0.24 |
| Female LGU | 0.34 | 0.08 | 1.56 | 0.17 |
| Female UGU | 0.94 | 0.44 | 2.03 | 0.88 |
| HPB | 2.22 | 1.17 | 4.22 | 0.02 |
| Joint | 1.18 | 0.62 | 2.25 | 0.62 |
| Lower GI | 2.70 | 1.46 | 4.99 | 0.00 |
| Major Vessel | 5.23 | 2.77 | 9.87 | 0.00 |
| Male GU | 0.13 | 0.02 | 0.97 | 0.05 |
| Muscle | 0.22 | 0.05 | 0.98 | 0.05 |
| Nasal | 0.20 | 0.06 | 0.73 | 0.02 |
| Neuro | 1.96 | 1.03 | 3.70 | 0.04 |
| Ocular | 0.03 | 0.01 | 0.08 | 0.00 |
| Oral | 0.49 | 0.20 | 1.18 | 0.11 |
| Other | 1.46 | 0.70 | 3.03 | 0.31 |
| Pharynx | 1.44 | 0.56 | 3.73 | 0.45 |
| Skin | 0.20 | 0.09 | 0.42 | 0.00 |
| Skull and Spine | 1.19 | 0.57 | 2.47 | 0.65 |
| Thoracic | 2.66 | 1.43 | 4.95 | 0.00 |
| Upper GI | 6.39 | 3.38 | 12.07 | 0.00 |
| Urological | 0.60 | 0.32 | 1.14 | 0.12 |
| Vascular | 1.51 | 0.80 | 2.85 | 0.21 |

**Supplementary table 2. Multivariable logistic regression analysis of death amongst patients undergoing elective surgery.** Patients with unspecified sex removed prior to analysis. Orthopaedic, Organ donation, Obstetrics and Cerebrovascular categories omitted as no SARS-CoV-2 patients in these groups. LGU; Lower genitourinary, UGU; Upper genitourinary, HPB; hepatopancreatobiliary, GI; gastrointestinal.

| **Variable** | **Odds ratio** | **Lower 95%CI** | **Upper 95%CI** | **p** |
| --- | --- | --- | --- | --- |
| COVID-19 | 5.65 | 5.35 | 5.95 | 0 |
| Female vs Male | 0.85 | 0.82 | 0.88 | 0 |
| Age | 1.03 | 1.03 | 1.03 | 0 |
| Cancer present | 1.01 | 0.97 | 1.05 | 0.54 |
| **Charlson co-morbidity score: vs.0** | | | | |
| Score 1 | 2.32 | 2.18 | 2.47 | 0 |
| Score 2 | 3.59 | 3.36 | 3.83 | 0 |
| Score 3 | 4.6 | 4.29 | 4.92 | 0 |
| Score 4 | 5.7 | 5.28 | 6.15 | 0 |
| Score 5 | 6.4 | 5.86 | 7 | 0 |
| Score >= 6 | 6.98 | 6.31 | 7.74 | 0 |
| **Index of multiple deprivation: vs quintile 5 (least deprived)** | | | | |
| Quintile 1 (most deprived) | 0.96 | 0.92 | 1.01 | 0.13 |
| Quintile 2 | 0.91 | 0.87 | 0.96 | 0 |
| Quintile 3 | 0.9 | 0.86 | 0.95 | 0 |
| Quintile 4 | 0.84 | 0.8 | 0.89 | 0 |
| **Procedure grouping: vs. Bone** | | | | |
| Breast | 0.49 | 0.23 | 1.04 | 0.06 |
| Cardiac | 1 | 0.93 | 1.07 | 0.94 |
| Cerebrovascular | 2.72 | 2.22 | 3.32 | 0 |
| Ear | 0.49 | 0.31 | 0.77 | 0 |
| Endocrine | 2.24 | 1.47 | 3.42 | 0 |
| Female LGU | 0.54 | 0.31 | 0.97 | 0.04 |
| Female UGU | 0.26 | 0.17 | 0.4 | 0 |
| HPB | 1.48 | 1.32 | 1.65 | 0 |
| Joint | 1.15 | 1.07 | 1.23 | 0 |
| Lower GI | 1.72 | 1.6 | 1.85 | 0 |
| Major Vessel | 2.54 | 2.28 | 2.84 | 0 |
| Male GU | 0.88 | 0.67 | 1.16 | 0.36 |
| Muscle | 1.01 | 0.8 | 1.28 | 0.93 |
| Nasal | 0.47 | 0.38 | 0.6 | 0 |
| Neuro | 4.01 | 3.73 | 4.32 | 0 |
| Organ Donation | 56.59 | 14.67 | 218.11 | 0 |
| Obstetrics | 0.65 | 0.21 | 2.01 | 0.45 |
| Ocular | 0.25 | 0.17 | 0.36 | 0 |
| Oral | 0.67 | 0.5 | 0.9 | 0.01 |
| Orthopaedics | 0.5 | 0.25 | 1.01 | 0.05 |
| Other | 3.14 | 2.78 | 3.56 | 0 |
| Pharynx | 1.88 | 1.28 | 2.75 | 0 |
| Skin | 0.91 | 0.83 | 0.99 | 0.03 |
| Skull and Spine | 1.49 | 1.26 | 1.76 | 0 |
| Thoracic | 3.91 | 3.66 | 4.18 | 0 |
| Upper GI | 3.19 | 2.92 | 3.5 | 0 |
| Urological | 1.02 | 0.92 | 1.12 | 0.75 |
| Vascular | 1.81 | 1.67 | 1.97 | 0 |

**Supplementary table 3. Multivariable logistic regression analysis of death amongst patients undergoing emergency surgery.** Patients with unspecified sex removed prior to analysis. LGU; Lower genitourinary, UGU; Upper genitourinary, HPB; hepatopancreatobiliary, GI; gastrointestinal.

|  | **No symptomatic codes** | **Symptomatic codes** |
| --- | --- | --- |
| **N** | 4171 | 7769 |
| **In-hospital death** | 344 (8.2%) | 2274 (29.3%) |
| **Length of hospital stay (days)** | 7 (2 to 18) | 17 (8 to 32) |

**Supplementary table 4. Number of patients with SARS-CoV-2 around the time of surgery, divided by the presence or absence of symptomatic codes.** Data are presented as n (%) or median (IQR). OPCS and ICD codes defining symptom status are listed in supplementary table 1.

| **Variable** | **Odds ratio** | **Lower 95%CI** | **Upper 95% CI** | **p** |
| --- | --- | --- | --- | --- |
| Presence of symptomatic codes | 3.73 | 3.28 | 4.25 | 0.00 |
| Female vs Male | 0.77 | 0.70 | 0.85 | 0.00 |
| Age | 1.04 | 1.03 | 1.04 | 0.00 |
| Cancer present | 1.18 | 1.04 | 1.33 | 0.00 |
| **Charlson co-morbidity score: vs.0** | | | | |
| Score 1 | 1.19 | 1.02 | 1.40 | 0.03 |
| Score 2 | 1.54 | 1.30 | 1.81 | 0.00 |
| Score 3 | 1.49 | 1.24 | 1.79 | 0.00 |
| Score 4 | 1.75 | 1.41 | 2.16 | 0.00 |
| Score 5 | 1.92 | 1.47 | 2.51 | 0.00 |
| Score >= 6 | 1.87 | 1.37 | 2.56 | 0.00 |
| **Index of multiple deprivation: vs quintile 5 (least deprived)** | | | | |
| Quintile 1 (most deprived) | 1.05 | 0.92 | 1.21 | 0.45 |
| Quintile 2 | 0.95 | 0.82 | 1.10 | 0.48 |
| Quintile 3 | 1.04 | 0.90 | 1.20 | 0.62 |
| Quintile 4 | 0.82 | 0.70 | 0.96 | 0.01 |
| **Admission category: vs Elective** | | | | |
| Emergency | 2.78 | 2.17 | 3.58 | 0.00 |
| Maternity | 0.13 | 0.01 | 1.29 | 0.08 |
| Other | 1.53 | 1.06 | 2.21 | 0.02 |
| **Procedure grouping: vs. Bone** | | | | |
| Cardiac | 0.74 | 0.57 | 0.96 | 0.02 |
| Cerebrovascular | 0.90 | 0.33 | 2.47 | 0.84 |
| Ear | 1.25 | 0.41 | 3.85 | 0.69 |
| Endocrine | 0.67 | 0.15 | 3.12 | 0.62 |
| Female LGU | 0.56 | 0.11 | 2.74 | 0.47 |
| Female UGU | 0.57 | 0.17 | 1.85 | 0.35 |
| HPB | 1.27 | 0.88 | 1.82 | 0.20 |
| Joint | 0.96 | 0.78 | 1.17 | 0.67 |
| Lower GI | 1.22 | 0.95 | 1.56 | 0.12 |
| Major Vessel | 1.12 | 0.74 | 1.69 | 0.59 |
| Male GU | 0.70 | 0.28 | 1.74 | 0.45 |
| Muscle | 2.40 | 1.18 | 4.89 | 0.02 |
| Nasal | 0.82 | 0.40 | 1.68 | 0.59 |
| Neuro | 1.27 | 0.98 | 1.64 | 0.07 |
| Obstetrics | 1.29 | 0.39 | 4.22 | 0.68 |
| Ocular | 0.60 | 0.20 | 1.76 | 0.35 |
| Oral | 1.32 | 0.51 | 3.42 | 0.57 |
| Orthopaedics | 0.39 | 0.04 | 3.44 | 0.40 |
| Other | 1.79 | 1.13 | 2.84 | 0.01 |
| Pharynx | 1.46 | 0.35 | 6.07 | 0.61 |
| Skin | 0.91 | 0.70 | 1.19 | 0.50 |
| Skull and Spine | 0.83 | 0.45 | 1.50 | 0.53 |
| Thoracic | 1.59 | 1.32 | 1.91 | 0.00 |
| Upper GI | 1.15 | 0.84 | 1.57 | 0.37 |
| Urological | 1.09 | 0.82 | 1.45 | 0.57 |
| Vascular | 1.79 | 1.40 | 2.28 | 0.00 |

**Supplementary table 5. Multivariable logistic regression analysis of death amongst patients with SARS-CoV-2 comparing those with symptomatic codes to those without.** Patients with unspecified sex removed prior to analysis. Breast omitted as no deaths in either group, Organ donation excluded as no symptomatic patients. LGU; Lower genitourinary, UGU; Upper genitourinary, HPB; hepatopancreatobiliary, GI; gastrointestinal.

| **SARS-CoV-2 PCR** | **ICD-10 code** | | **Number** |
| --- | --- | --- | --- |
| - | Absent | | 86052 |
| - | Present | | 289 |
| + | Present | | 441 |
| + | Absent | | 84 |
| **Incidence based on:** | | | |
| **ICD-10 codes only** | | 730 (0.8%) | |
| **SARS-CoV-2 PCR only** | | 525 (0.6%) | |
| **ICD-10 codes**  **and**  **SARS-CoV-2 PCR** | | 814 (0.9%) | |

**Supplementary table 6. Data drawn from PEDW for all patients undergoing surgery in Wales.** SARS-CoV-2 PCR; Polymerase Chain Reaction test for SARS-CoV-2. PCR test data was captured during the same time window as ICD-10 codes to estimate the incidence of SARS-CoV-2 recorded by different methods. ICD-10 codes are assigned based on clinical diagnosis. -; no positive SARS-CoV-2 PCR during time window, +; one or more positive during time window.

| **OPCS** | **Description** | **Reason** |
| --- | --- | --- |
| **L91.X** | Open insertion of central venous catheter | Often used for bedside central line insertion |
| **L99.X** | Other therapeutic transluminal operations. | Often used for bedside central line insertion |
| **L71.4** | Percutaneous transluminal cannulation of artery | Often used for bedside arterial line insertion |

**Supplementary table 7. OPCS Codes excluded from the definition of surgery, with reasons**
